# Comparative Analysis of ChatGPT’s Diagnostic Performance with Radiologists Using Real-World Radiology Reports of Brain Tumors

**DOI:** 10.1101/2023.10.27.23297585

**Authors:** Yasuhito Mitsuyama, Hiroyuki Tatekawa, Hirotaka Takita, Fumi Sasaki, Akane Tashiro, Satoshi Oue, Shannon L Walston, Yukio Miki, Daiju Ueda

## Abstract

**Background:** Large Language Models like Chat Generative Pre-trained Transformer (ChatGPT) have demonstrated potential for differential diagnosis in radiology. Previous studies investigating this potential primarily utilized quizzes from academic journals, which may not accurately represent real-world clinical scenarios.

**Purpose:** This study aimed to assess the diagnostic capabilities of ChatGPT using actual clinical radiology reports of brain tumors and compare its performance with that of neuroradiologists and general radiologists.

**Methods:** We consecutively collected brain MRI reports from preoperative brain tumor patients at Osaka Metropolitan University Hospital, taken from January to December 2021. ChatGPT and five radiologists were presented with the same findings from the reports and asked to suggest differential and final diagnoses. The pathological diagnosis of the excised tumor served as the ground truth. Chi-square tests and Fisher’s exact test were used for statistical analysis.

**Results:** In a study analyzing 99 radiological reports, ChatGPT achieved a final diagnostic accuracy of 75% (95% CI: 66, 83%), while radiologists’ accuracy ranged from 64% to 82%. ChatGPT’s final diagnostic accuracy using reports from neuroradiologists was higher at 82% (95% CI: 71, 89%), compared to 52% (95% CI: 33, 71%) using those from general radiologists with a p-value of 0.012. In the realm of differential diagnoses, ChatGPT’s accuracy was 95% (95% CI: 91, 99%), while radiologists’ fell between 74% and 88%. Notably, for these differential diagnoses, ChatGPT’s accuracy remained consistent whether reports were from neuroradiologists (96%, 95% CI: 89, 99%) or general radiologists (91%, 95% CI: 73, 98%) with a p-value of 0.33.

**Conclusion:** ChatGPT exhibited good diagnostic capability, comparable to neuroradiologists in differentiating brain tumors from MRI reports. ChatGPT can be a second opinion for neuroradiologists on final diagnoses and a guidance tool for general radiologists and residents, especially for understanding diagnostic cues and handling challenging cases.

**Summary:** This study evaluated ChatGPT’s diagnostic capabilities using real-world clinical MRI reports from brain tumor cases, revealing that its accuracy in interpreting brain tumors from MRI findings is competitive with radiologists.

**Key results:**

- ChatGPT demonstrated a diagnostic accuracy rate of 75% for final diagnoses based on preoperative MRI findings from 99 brain tumor cases, competing favorably with five radiologists whose accuracies ranged between 64% and 82%. For differential diagnoses, ChatGPT achieved a remarkable 95% accuracy, outperforming several of the radiologists.
- Radiology reports from neuroradiologists and general radiologists showed varying accuracy when input into ChatGPT. Reports from neuroradiologists resulted in higher diagnostic accuracy for final diagnoses, while there was no difference in accuracy for differential diagnoses between neuroradiologists and general radiologists.

## Introduction

The emergence and subsequent advancements of Large Language Models (LLMs) like Chat Generative Pre-trained Transformer (ChatGPT) have recently dominated global technology discourse (1). These models represent a new frontier in artificial intelligence, using machine learning techniques to process and generate human language in a way that rivals human-level complexity and nuance. The rapid evolution and widespread impact of LLMs has become a global phenomenon, prompting discussions on their potential applications and implications (2–5).

Within the realm of LLMs, the GPT series, in particular, has gained significant attention. Many applications have been explored within the field of radiology (6–21). Among these, the potential of GPT to assist in diagnosis from image findings is noteworthy (18–20) because such capabilities could complement the essential aspects of daily clinical practice and education. Two studies show the potential of ChatGPT based on GPT-4 to generate differential diagnosis in the field of neuroradiology (19,20). One study utilizes the “Case of the Week’’ from the American Journal of Neuroradiology (19) and the other study utilizes ’’Freiburg Neuropathology Case Conference’’ cases from the Clinical Neuroradiology journal (20).

Although these pioneering investigations suggest that ChatGPT could play an important role in radiological diagnosis, there are no studies reporting evaluation using real-world radiology reports. Unlike quizzes (19,20), which tend to present carefully curated, typical cases and are created by individuals already aware of the correct diagnosis, real-world radiology reports may contain less structured and more diverse information. This difference might lead to biased evaluations that do not reflect the complex nature of clinical radiology (22,23).

To address this gap, our study examines the diagnostic abilities of ChatGPT using only real-world clinical radiology reports. We zeroed in on MRI reports pertaining to brain tumors, given the pivotal role radiological reports play in determining treatment routes such as surgery, medication, or monitoring; and that pathological outcomes offer a definitive ground truth for brain tumors (24). We compare the performance of ChatGPT with that of neuroradiologists and general radiologists, aiming to provide a more comprehensive view. Through this investigation, we aim to uncover the capabilities and potential limitations of ChatGPT as a diagnostic tool in a real-world clinical setting.

## Methods

### Study design

In our daily clinical practice, thinking through differential and final diagnoses can be challenging and time-consuming. If ChatGPT can excel in this diagnostic process, it indicates potential value in clinical scenarios. To test this, we presented ChatGPT with imaging findings from our real reports and asked it to suggest differential and final diagnoses. For a fair comparison, we also presented the same image findings (not images) to radiologists and requested differential diagnoses and a final diagnosis. The protocol of this study was reviewed and approved (approval no. 2023-015) by the Ethical Committee of the Osaka Metropolitan University Graduate School of Medicine. This study was conducted in accordance with the Declaration of Helsinki. The requirement for informed consent was waived because the radiology reports had been acquired during daily clinical practice. The design of this study is based on the Standards for Reporting of Diagnostic Accuracy Studies (STARD) guideline (25).

### Data collection

In this study, we collected brain MRI image findings of preoperative brain tumors from radiological reports taken at Osaka Metropolitan University Hospital from January 2021 to December 2021. Out of the collected cases, we excluded those that involved reoperation. These imaging findings were subsequently verified by two radiologists (7 years of experience; 4 years of experience). When a diagnosis was described in the imaging findings, it was also removed to avoid data leakage. Any descriptions related to previous imaging findings and unrelated image descriptors (such as ’figure 1’), were deleted. The report writer (neuroradiologist or general radiologist) was noted. These simplified findings were subsequently verified by the same two radiologists.

**Figure 1.**
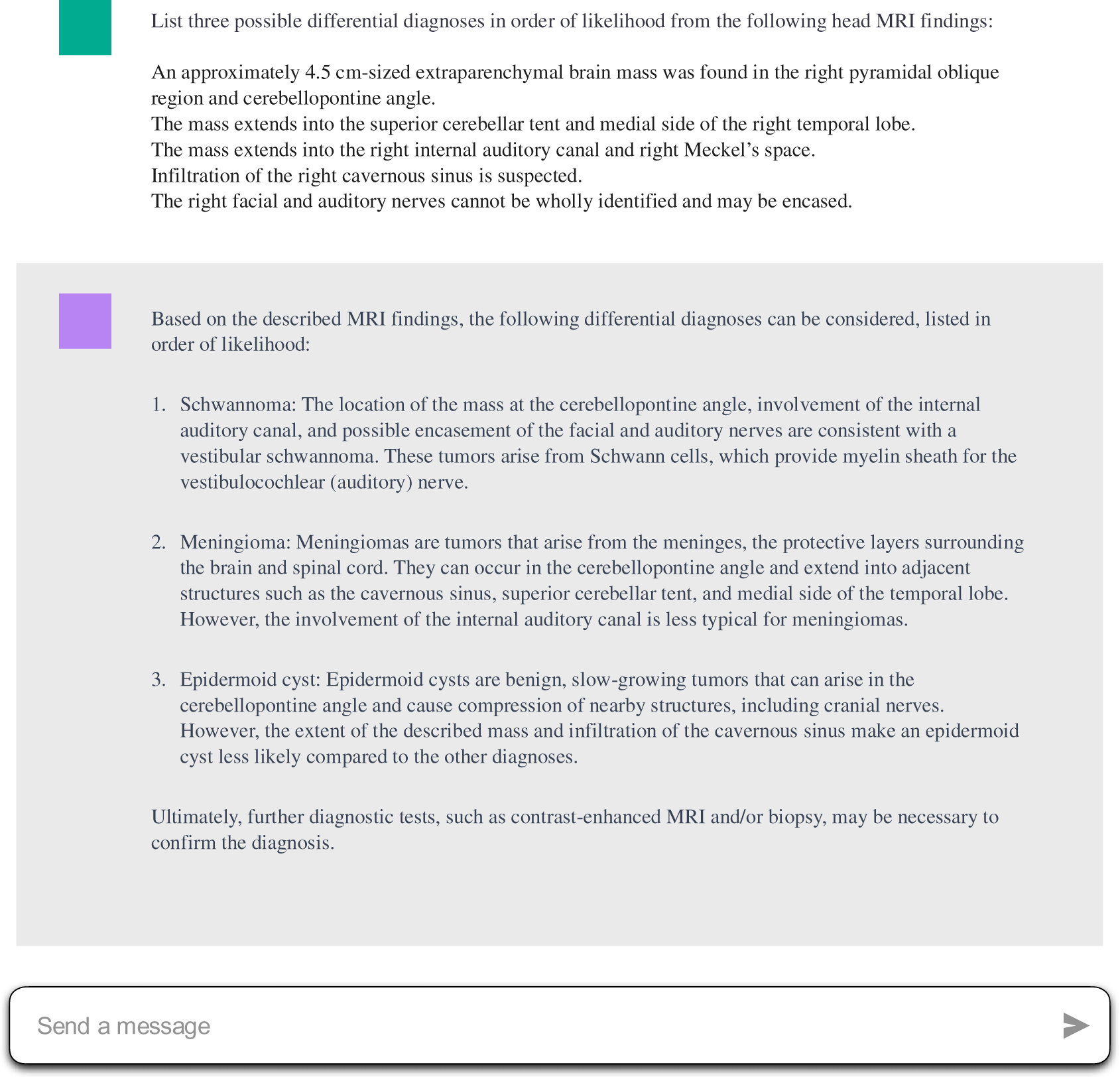
Examples of interface with ChatGPT. These are input texts (simplified MRI imaging findings) to ChatGPT and output texts generated by ChatGPT. The diagnosis listed highest among the three differential diagnoses was determined to be the final diagnosis.

### Input and output procedure for ChatGPT

We input the following premise into ChatGPT based on the GPT-4 architecture (May 24 version; OpenAI, California, USA; https://chat.openai.com/): *List three possible differential diagnoses in order of likelihood from the following head MRI findings.* Then, we input the imaging findings and received three differential diagnoses from ChatGPT. The diagnosis listed highest among the three differential diagnoses was determined to be the final diagnosis. An example of the input to ChatGPT and the output of ChatGPT is shown in Figure 1.

### Radiologist reading test

We provided the same image findings input into ChatGPT to five radiologists, including two neuroradiologists (Radiologist A; 13 years of experience, Radiologist B; 8 years of experience) and three general radiologists (Radiologist C; 4 years of experience, Radiologist D; 3 years of experience, Radiologist E; 2 years of experience). They read these findings and provided three differential diagnoses including one final diagnosis.

### Output evaluation

We utilized the pathological diagnosis of the tumor that was excised in neurosurgery as the ground truth. Two radiologists (7 years of experience; 4 years of experience) confirmed whether the differential diagnoses and final diagnosis suggested by ChatGPT and the actual ground truth were the same. We introduced this process of confirmation because the ground truth diagnosis may use alternative words or phrasing. Likewise, the radiologists’ final and differential diagnoses were also reviewed for accuracy by the same two radiologists.

### Statistical analysis

We computed the accuracy rates of both the differential and final diagnoses made by ChatGPT and those of the five radiologists. To compare the diagnostic accuracy of the differential and final diagnoses between ChatGPT and each radiologist, we conducted Chi-square tests. Additionally, we calculated these accuracy rates separately for when the reporter was a neuroradiologist and when the reporter was a general radiologist to examine how the quality of input (image findings) affects the diagnoses both by ChatGPT and radiologists. Moreover, Fisher’s exact test was performed to compare the diagnostic accuracy, both of ChatGPT and the five radiologists, resulting from the reports by neuroradiologist or general radiologist reporters. All analyses were performed using R (version 4.0.0, 2020; R Foundation for Statistical Computing; https://R-project.org).

## Results

A total of 99 radiological reports were included in this research after excluding 69 reports because of previous operation. A flowchart of data collection is shown in Figure 2. The final diagnostic accuracy rates for ChatGPT and Radiologists A, B, C, D, and E were 75% (95% CI: 66, 83%), 69% (95% CI: 60, 78%), 82% (95% CI: 74, 89%), 66% (95% CI: 56, 75%), 75% (95% CI: 66, 83%), and 64% (95% CI: 54, 73%), respectively. The chi-square test showed significant differences between the correct diagnosis rates of ChatGPT and Radiologists B, C, D, and E (p-values; Radiologist B: <0.001, Radiologist C: <0.001, Radiologist D: <0.001, Radiologist E: 0.002) but not Radiologist A (p-values: 0.067). The correct differential diagnosis rates for ChatGPT and Radiologists A, B, C, D, and E were 95% (95% CI: 91, 99%), 87% (95% CI: 80, 94%), 88% (95% CI: 81, 94%), 78% (95% CI: 70, 86%), 82% (95% CI: 74, 89%), and 74% (95% CI: 65, 82%), respectively. The chi-square test showed significant differences between the correct diagnosis rates of ChatGPT and Radiologists B, C, and E (p-values; Radiologist B: <0.001, Radiologist C: <0.001, Radiologist E: <0.001) but not Radiologists A and D (p-values; Radiologist A: 0.25, Radiologist D: 0.48).

**Figure 2.**
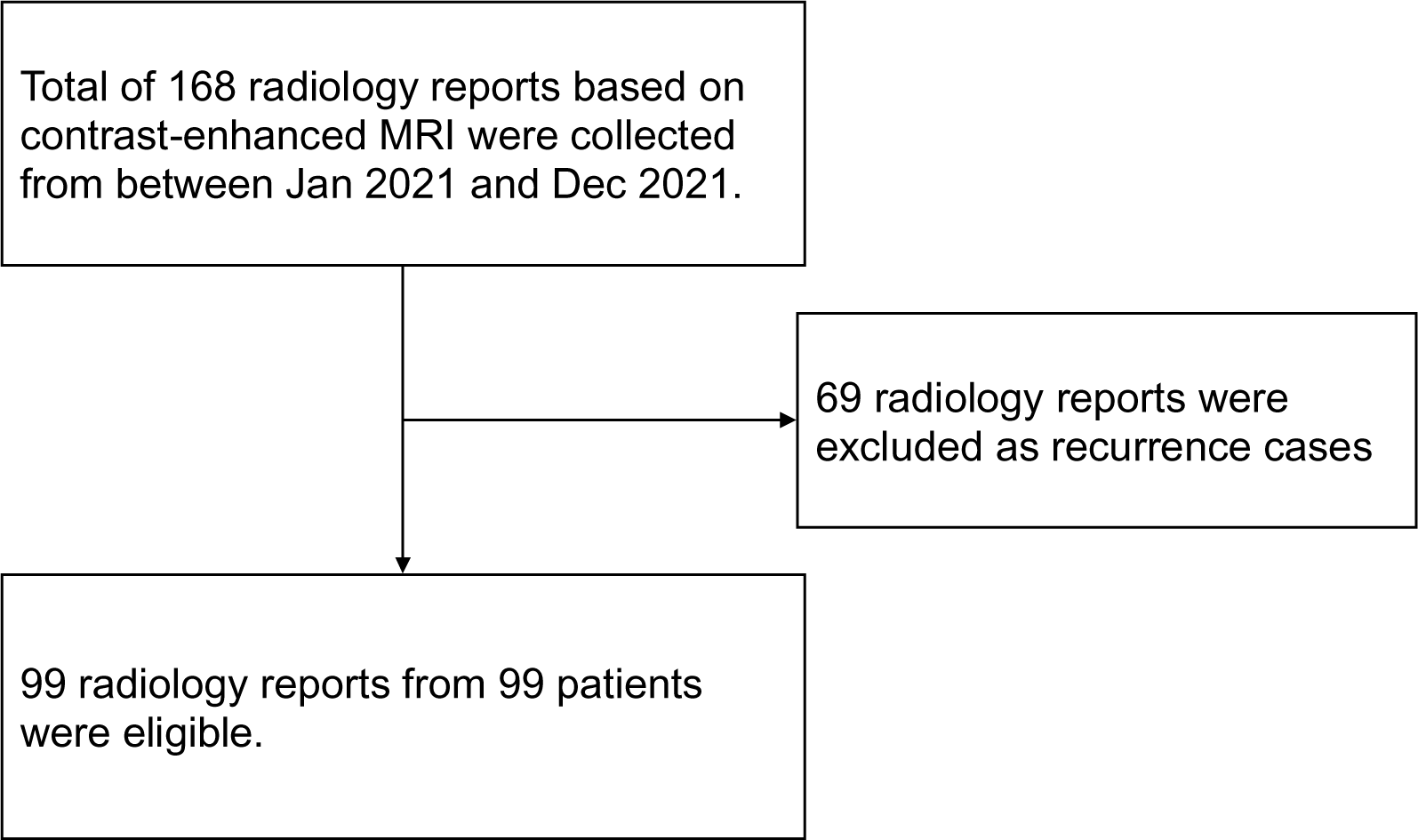
Flowchart of data collection. This is the data collection flowchart.

As for the final diagnosis, with the findings from reports written by neuroradiologists, the correct diagnosis rates by ChatGPT and Radiologists A, B, C, D, and E were 82% (95% CI: 71, 89%), 71% (95% CI: 60, 80%), 82% (95% CI: 71, 89%), 67% (95% CI: 56, 77%), 79% (95% CI: 69, 87%), and 66% (95% CI: 55, 75%), respectively. With the findings from reports written by general radiologists, the correct final diagnosis rates for the final diagnoses by ChatGPT and Radiologist A, B, C, D, and E were 52% (95% CI: 33, 71%), 61% (95% CI: 41, 78%), 83% (95% CI: 63, 93%), 61% (95% CI: 41, 78%), 61% (95% CI: 41, 78%), and 57% (95% CI: 37, 74%), respectively. ChatGPT performed statistically better with the neuroradiologists’ reports than with the general radiologists’ reports (p-value: 0.012). Radiologists do not show statistical differences between reading reports written by neuroradiologists and those written by general radiologists (p-values; Radiologist A: 0.44, Radiologist B: >0.99, Radiologist C: 0.62, Radiologist D: 0.10, Radiologist E: 0.46).

As for the differential diagnoses determined using the findings from reports by neuroradiologists, the correct diagnosis rates by ChatGPT and Radiologists A, B, C, D, and E were 96% (95% CI: 89, 99%), 89% (95% CI: 81, 95%), 88% (95% CI: 79, 94%), 78% (95% CI: 67, 86%), 83% (95% CI: 73, 90%), and 75% (95% CI: 64, 83%), respectively. Using the findings from reports by general radiologists, the correct diagnosis rates by ChatGPT and Radiologist A, B, C, D, and E were 91% (95% CI: 73, 98%), 78% (95% CI: 58, 90%), 87% (95% CI: 68, 95%), 78% (95% CI: 58, 90%), 78% (95% CI: 58, 90%), and 70% (95% CI: 49, 84%), respectively. ChatGPT does not have significantly different performance using either the neuroradiologists’ report or the general radiologists’ report (p-values: 0.33). Radiologists also do not show significant differences when using either the neuroradiologists’ report or the general radiologists’ report (p-values; Radiologist A: 0.17, Radiologist B: >0.99, Radiologist C: >0.99, Radiologist D: 0.76, Radiologist E: 0.60). The accuracy rates of the ChatGPT and the five radiologists in the final and differential diagnoses are shown in Figure 3.

**Figure 3.**
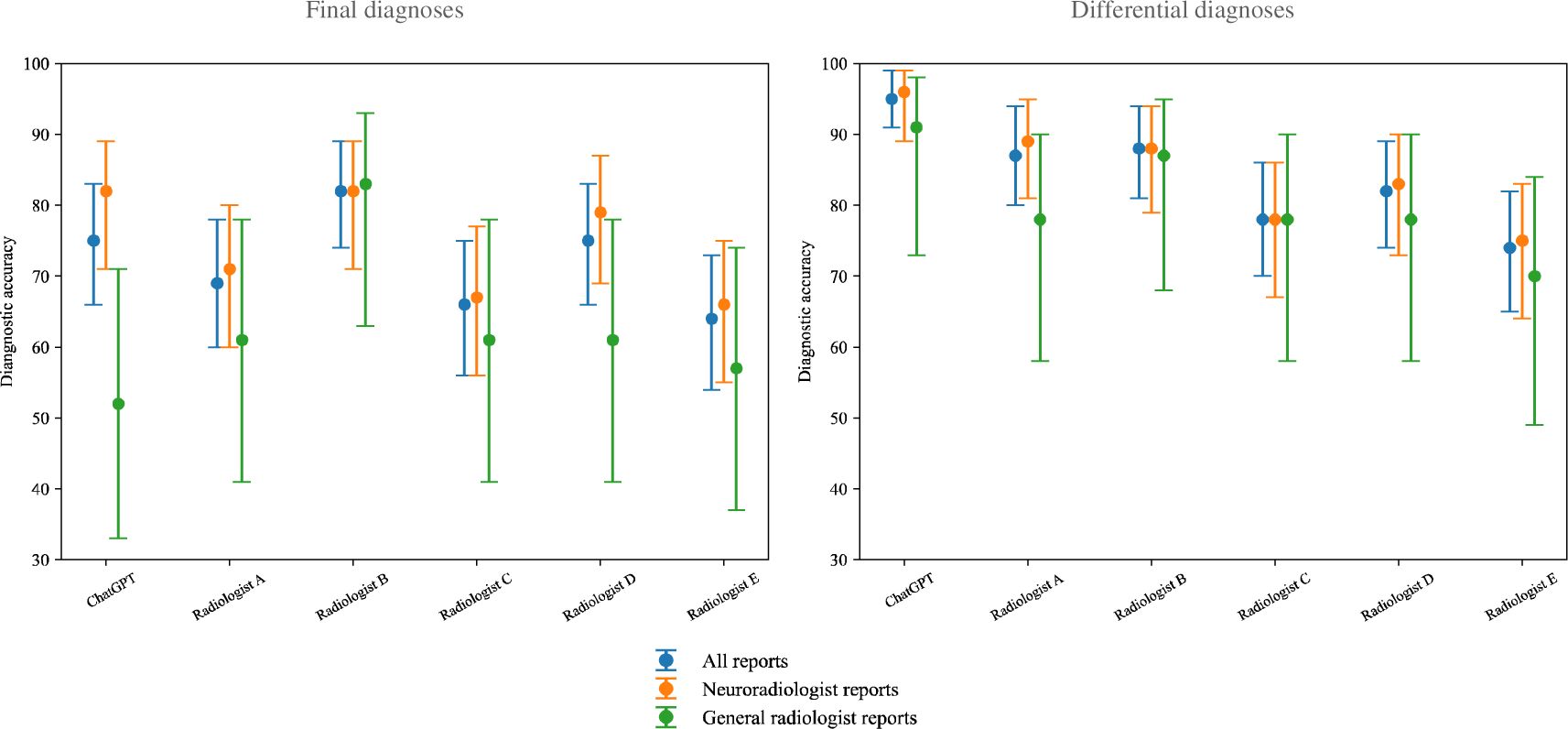
Accuracy of Chatgpt and Radiologists. The bar-plots represent the accuracy of ChatGPT and radiologists for the final and differential diagnoses, respectively.

## Discussion

ChatGPT and five radiologists were presented with preoperative brain MRI findings from 99 cases and asked to list differential and final diagnoses. For final diagnoses, ChatGPT’s accuracy was 75% (95% CI: 66, 83%). In comparison, Radiologists A through E had accuracies of 69% (95% CI: 60, 78%), 82% (95% CI: 74, 89%), 66% (95% CI: 56, 75%), 75% (95% CI: 66, 83%), and 64% (95% CI: 54, 73%), respectively. For differential diagnoses, ChatGPT achieved 95% (95% CI: 91, 99%) accuracy, while the radiologists’ accuracies ranged from 74% (95% CI: 65, 82%) to 88% (95% CI: 81, 94%). In the final diagnoses, ChatGPT showed an accuracy of 82% (95% CI: 71, 89%) with reports from neuroradiologists, compared to 52% (95% CI: 33, 71%) with those from general radiologists, a statistically significant difference (p-value: 0.012). On the other hand, ChatGPT’s differential diagnostic accuracy was 96% (95% CI: 89, 99%) with reports from neuroradiologists and 91% (95% CI: 73, 98%) with reports from general radiologists, not a statistically significant difference (p-value: 0.33).

This study is the first attempt to evaluate ChatGPT’s ability to interpret actual clinical radiology reports, rather than from settings like image diagnosis quizzes. The majority of previous research (6–12,17–21) suggested the utility of ChatGPT in diagnostics, but these relied heavily on hypothetical environments such as quizzes from academic journals or examination questions (26). This approach can lead to a cognitive bias since the individuals formulating the imaging findings or exam questions also possess the answers. In these simulated scenarios, there’s also a propensity to leave out minor findings. Such minor findings, while often deemed insignificant in an experimental setup, are frequently encountered in real-world clinical practice and can have implications on diagnosis. In contrast, our study deviates from this previous methodology by using actual clinical findings, generated in a state of diagnostic uncertainty. This approach facilitates a more robust and practical evaluation of ChatGPT’s accuracy, keeping in mind its potential applications in real-world clinical settings.

When reports created by neuroradiologists and general radiologists were input into ChatGPT, there was a notable difference in ChatGPT’s diagnostic accuracy. Specifically, for the final diagnosis, using reports from the neuroradiologists yielded higher accuracy than using those from general radiologists. However, for differential diagnoses, there was no difference in accuracy, regardless of whether the report was from a neuroradiologist or a general radiologist. Neuroradiologists, due to their experience and specialized knowledge, are more likely to include comprehensive, detailed information necessary for a final diagnosis in their reports (27–29). Such high-quality reports likely enhanced ChatGPT’s accuracy for final diagnoses. Conversely, ChatGPT possesses the ability to provide accurate differential diagnoses even with the general radiologists report because they can capture certain information crucial for a diagnosis. From these findings, a beneficial application of ChatGPT in clinical and educational settings is for neuroradiologists to use it as a second opinion to assist with final and differential diagnoses. For general radiologists, ChatGPT can be particularly useful for understanding diagnostic cues and learning about differential diagnoses, which can sometimes be time consuming. When general radiologists encounter complex or unfamiliar cases, consulting ChatGPT could guide their diagnostic direction. Of course, any advice or suggestions from ChatGPT should be considered as just one of many references. General radiologists should prioritize consultation with experts when determining the final diagnosis.

There are several limitations. This study only used the wording of actual clinical radiology reports and did not evaluate the effect of including other information such as patient history and the image itself, meaning the radiologists’ performance might not match their real-world diagnostic abilities. As only data from a single institution was used, the findings and cases might be biased, and validation using data from multiple institutions and larger data sets is desired. We did not assess MRI reports for diseases other than brain tumors.

ChatGPT has showcased a great diagnostic ability, demonstrating performance comparable to that of neuroradiologists in the task of diagnosing brain tumors from MRI reports. The implications of these findings are far-reaching, suggesting potential real-world utility, particularly in the generation of differential diagnoses for general radiologists in a clinical setting. The encouraging results of this study invite further evaluations of the LLM’s accuracy across a myriad of medical fields and imaging modalities. The end goal of such exploration is to pave the way for the development of more versatile, reliable, and powerful tools for healthcare.

**Table 1:** Demographics of Radiologists.

|  | Experience (years) | Sex |
| --- | --- | --- |
| Neuroradiologists |  |  |
| Radiologist A | 12 | M |
| Radiologist B | 9 | M |
| General radiologists |  |  |
| Radiologist C | 6 | F |
| Radiologist D | 5 | F |
| Radiologist E | 4 | M |

**Table 2:** Demographics of Brain MRI Cases.

|  | Number |
| --- | --- |
| Examination | 99 |
| Male | 35 |
| Female | 64 |
| Mean (sd) age | 53 ± 17 |
| Pathology |  |
| Meningioma | 34 |
| Pituitary adenoma | 17 |
| Schwannoma | 12 |
| Angioma | 5 |
| Craniopharyngioma | 4 |
| Hemangioblastoma | 4 |
| High grade glioma | 10 |
| Glioblastoma | 4 |
| Anaplastic astrocytoma | 2 |
| Anaplastic oligodendroglioma | 2 |
| Unknown | 2 |
| Low grade glioma | 3 |
| Diffuse astrocytoma | 1 |
| Unknown | 2 |
| Epidermal cyst | 2 |
| Sarcoma | 2 |
| Arachnoid | 1 |
| Chordoma | 1 |
| Lymphoma | 1 |
| Metastatic tumor | 1 |
| Rathke's cleft cyst | 1 |
| Central neurocytoma | 1 |
| Reporter type |  |
| Neuroradiologist | 76 |
| General radiologist | 23 |

**Table 3:** Results for ChatGPT and Radiologists.

|  | Accuracy (%) (95%CI) | Chi-square statistic | p-value |
| --- | --- | --- | --- |
| Final diagnosis |  |  |  |
| ChatGPT | 75 (66–83) | NA | NA |
| Radiologist A | 69 (60–78) | 3.4 | 0.067 |
| Radiologist B | 82 (74–89) | 12.8 | <0.001 |
| Radiologist C | 66 (56–75) | 11.3 | <0.001 |
| Radiologist D | 75 (66–83) | 10.9 | <0.001 |
| Radiologist E | 64 (54–73) | 9.5 | 0.002 |
| Differential diagnosis |  |  |  |
| ChatGPT | 95 (91–99) | NA | NA |
| Radiologist A | 87 (80–94) | 1.3 | 0.25 |
| Radiologist B | 88 (81–94) | 7.1 | 0.008 |
| Radiologist C | 78 (70–86) | 14.0 | <0.001 |
| Radiologist D | 82 (74–89) | 0.49 | 0.48 |
| Radiologist E | 74 (65–82) | 11.0 | <0.001 |

**Table 4:** Results for ChatGPT and Radiologists by reporter.

|  | Accuracy (%) (95%CI) | Accuracy (%) (95%CI) | Fisher's exact test |
| --- | --- | --- | --- |
|  | Neuroradiologist | General radiologist | p-value |
| Final diagnosis |  |  |  |
| ChatGPT | 82 (71–89) | 52 (33–71) | **0.012** |
| Radiologist A | 71 (60–80) | 61 (41–78) | 0.44 |
| Radiologist B | 82 (71–89) | 83 (63–93) | >0.99 |
| Radiologist C | 67 (56–77) | 61 (41–78) | 0.62 |
| Radiologist D | 79 (69–87) | 61 (41–78) | 0.10 |
| Radiologist E | 66 (55–75) | 57 (37–74) | 0.46 |
| Differential diagnosis |  |  |  |
| ChatGPT | 96 (89–99) | 91 (73–98) | 0.33 |
| Radiologist A | 89 (81–95) | 78 (58–90) | 0.17 |
| Radiologist B | 88 (79–94) | 87 (68–95) | >0.99 |
| Radiologist C | 78 (67–86) | 78 (58–90) | >0.99 |
| Radiologist D | 83 (73–90) | 78 (58–90) | 0.76 |
| Radiologist E | 75 (64–83) | 70 (49–84) | 0.60 |

## Data Availability

All data produced in the present study are available upon reasonable request to the authors

## Acknowledgement

We have used ChatGPT to generate a portion of the manuscript, but the output was confirmed by the authors.

